# Ambient temperature and impacts on preterm birth: A storyline approach

**DOI:** 10.64898/2026.10.07.26364935

**Authors:** Ricardo Berenguer Navarro, Gabriele Messori, Jette Möller, Ilias Pechlivanidis, Denica Bozhinova, Katharina Klehmet, Elena Raffetti

## Abstract

**Background:** Non-optimal air temperature increases the risk of preterm birth, but how temperature-related risks may translate into future population burden remains uncertain. We aimed to estimate the association between temperature and preterm birth in Sweden and assess how preterm birth burden may change under plausible future temperature conditions.

**Methods:** We conducted a nationwide population-based cohort study of singleton births in Sweden from 1973 to 2018. Weekly mean temperature was assigned at the parish level according to maternal residence. Cox proportional hazards models with a distributed lag nonlinear model were used to estimate nonlinear and delayed associations between temperature and preterm birth. The estimated exposure–response relationship and baseline hazards were then incorporated into an individual-based microsimulation of gestational progression under projected temperatures for 2025–2055. Future projections were examined across multiple climate models under SSP1-2.6 and SSP3-7.0 scenarios. Outcomes included the annual number and cumulative incidence of preterm births and the temperature-attributable fraction.

**Results:** Among 4,338,825 singleton births, 211,397 (4.9%) were preterm. Temperature–preterm birth associations were modest overall. The clearest increase in hazard was observed following exposure to very low temperatures, particularly −10°C at short lags. Higher temperatures showed no clear increase in risk. From 2025 to 2055, the annual cumulative incidence of preterm birth and the temperature-attributable fraction was expected to remain broadly stable, with limited differences between emissions scenarios. In contrast, the absolute number of preterm births was expected to over the projection period, primarily reflecting increases in the projected number of pregnancies rather than substantial changes in temperature-associated risk.

**Conclusions:** Very low temperatures were associated with a modest short-term increase in preterm birth risk in Sweden, hot temperatures showed little evidence of increased risk. Applying these associations to future climate conditions suggested limited changes in preterm birth incidence or the proportion attributable to temperature through 2055. However, the absolute burden of preterm birth may increase as the population at risk grows. This highlights the importance of considering demographic change alongside climate projections when assessing future maternal and neonatal health burden.

## Introduction

Temperature-related risks during pregnancy are not limited to heat. In high-latitude countries such as Sweden, cold exposure remains relevant even as average temperatures rise and heat extremes become more frequent. Epidemiological evidence suggests that exposure to both cold and hot ambient temperatures is associated with an increased risk of preterm birth (Dalugoda et al. 2024, Lakhoo et al. 2025, Girgin et al. 2025, Wang et al. 2025, Ge et al. 2026). Increased cardiovascular demand and metabolic requirements, together with altered thermoregulation, may heighten susceptibility to temperature-related stress during pregnancy (Raffetti et al. 2026).

Preterm birth, defined as a live birth before 37 completed weeks of gestation, is a major public health concern. Children born preterm face increased risks of respiratory disease, neurodevelopmental impairment, and long-term morbidity, placing a substantial burden on families and health-care systems (Undela et al. 2019, Ohuma et al. 2023, Inder et al. 2023).

Climate change is raising temperatures worldwide, with Europe warming at approximately twice the global average (Copernicus Climate Change Service and WMO, 2025; van Daalen et al., 2024). These changes are having substantial impacts on urban environments and may disproportionately affect vulnerable populations (Mishra et al., 2015; Macintyre et al., 2018; Tuholske et al., 2021). Although cold extremes are expected to become less frequent, they will continue to occur, particularly at high latitudes. Estimating the future burden of temperature-related preterm birth requires consideration of the full temperature distribution rather than heat exposure alone. Retrospective studies have long informed public health by identifying associations between environmental exposures and health outcomes, but they cannot alone estimate risks under future conditions. Simulation models and climate projections offer a forward-looking perspective, but their estimates are sensitive to assumptions about exposure-response relationships, demographic change, adaptation, and the magnitude, timing, and spatial distribution of future temperature change (Gasparrini, 2017, Rai et al. 2022, Weber et al. 2023, Cordiner et al. 2024, Messori et al. 2026).

Climate storylines complement these methods by providing a nonprobabilistic framework for examining plausible, context-specific future risks. They use climate projections to develop physically plausible and internally consistent scenarios of how specific climatic conditions or extreme events might unfold (Hazeleger 2015; Shepherd et al. 2018). In practice, storylines connect projected climatic conditions with information about who will be exposed, how vulnerable they are, and their capacity to adapt. By linking climate hazards to population-health outcomes, storylines can clarify potential consequences and support adaptation planning when event probabilities are uncertain (Zscheischler et al. 2018; Rusca et al. 2021, Rusca et al. 2024). Any resulting estimates are conditional on the scenarios examined and should not be interpreted as probabilistic forecasts.

In this study, we extend the application of climate storylines to population health. We aim to characterise birth risk under plausible future temperature conditions in Sweden leveraging climate projections, retrospective epidemiological analysis, and microsimulation. This integrated approach accounts for both climatic and population-level dynamics. This addresses three research questions: (1) how is ambient temperature associated with the risk of preterm birth in Sweden under historical conditions; (2) how might preterm birth change under projected future temperatures; and (3) how can a storyline-based framework be used to incorporate population and broader social dynamics into projections of future temperature-related preterm birth? To this end, we first estimate the historical temperature–preterm birth relationship using retrospective data and then incorporate this relationship into a simulation framework under projected future temperatures. The storyline approach then combines climate projections with plausible population and social changes. This allows the future burden of temperature-related preterm birth to be assessed across different scenarios.

## Methods Study design

We conducted a population-based cohort study of pregnant women from 1973 to 2018 using Swedish national registry data. The study population included all singleton births in Sweden during the study period. We excluded records with missing information on temperature exposure and other key covariates. Multiple births from the same mother within 22 weeks were excluded. The exposure was defined as mean weekly temperature (°C) assigned at parish level. For every week of pregnancy, weekly mean temperature was assigned as a time-varying exposure. The primary outcome was time to delivery, with gestational age measured in weeks. Deliveries were classified as preterm birth (≥22 and <37 completed weeks of gestation) or term birth (≥37 completed weeks of gestation).

## Data sources

Information on births and pregnancy characteristics was obtained from the Swedish Medical Birth Register. The register provided data on gestational age, date of birth, maternal characteristics, and other pregnancy-related information. Individual-level demographic and residential information was obtained from population registers maintained by Statistics Sweden, including information on place of residence and socioeconomic characteristics. Individual-level data from the different registers were linked using each resident’s unique personal identification number.

Daily mean temperature data were obtained from the Swedish Meteorological and Hydrological Institute (SMHI) at a spatial resolution of 2.5 km x 2.5 km for the periods 1961-2018 from the SMHIGridCLIM observational dataset (Andersson et al., 2021; Andersson et al., 2026, in review)..Future climate projections from 5 CMIP6 global climate models for the period of 2015-2100 were downscaled using the HCLIM43-ALADIN regional climate model as part of the CMIP6-based EURO-CORDEX experiment (Nikulin et al. 2026)The climate data was further bias-adjusted and downscaled from the original 12 km horizontal resolution to the 2.5 km of the SMHIGridCLIM grid using 1981-2010 as reference period with the MIdAS (MultI-scale bias AdjuStment) method (Berg et al., 2022).

We averaged the daily temperatures in each parish for every gestational week. Each pregnancy was then assigned the weekly mean temperature for the parish of residence from estimated conception until delivery.

### Statistical analysis

### Retrospective analysis

We used Cox proportional hazards models to estimate the association between temperature and time to delivery, with gestational week as the time scale and preterm birth defined as the binary event indicator. Temperature was incorporated through a distributed lag nonlinear model cross-basis, allowing for nonlinear and delayed associations (Gasparrini et al. 2010). Natural cubic splines were used for the exposure–response function and B-splines for the lag–response function. Models were adjusted for maternal age, seasonality, income level, and previous maternal hospitalization, identified using a prespecified list of diagnoses. Sensitivity analyses compared the fully adjusted model with 6 degrees of freedom for seasonality against models using alternative seasonal degrees of freedom, an adjusted model without seasonality, and an unadjusted model.

Let *T*_*i*_ denote the gestational duration for individual i. The hazard of delivery at gestational week t was modelled as:

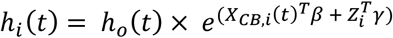

Where *h*_*o*_*(t)* represent the baseline hazard. The term *X*_*CB,i*_(*t*)represents the cross-basis matrix for the exposure of interest, allowing simultaneous modeling of the exposure–response relationship and lagged effects over time. The vector *β*contains the corresponding coefficients for the cross-basis terms. The vector *Z*_*i*_includes additional covariates, with associated regression coefficients γ.

We incorporated the estimated model coefficients into an individual-based microsimulation of gestational progression under projected temperature exposures (Brunn et al. 2026). The microsimulation translated the exposure–lag–response association into a probability of delivery for each week of gestation.

The estimated regression coefficient and the baseline hazard were extracted from the Cox Proportional Hazards model and used to calculate week-specific hazards of delivery for each simulated pregnancy. Temperature projections from the Swedish Meteorology and Hydrology institute were assigned according to the selected heatwave scenario, and the corresponding exposure history were transformed through the DLNM function to generate gestational week specific hazard estimates.

The microsimulation proceeded in discrete weekly steps from conception. At each week *t* pregnancies remained at risk until transitioning to the absorbing states of preterm birth or term birth. The conditional probabilities of delivery at each time point was derived from the estimated hazard function as:

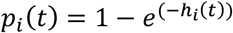

For each simulated individual and gestational week, a random draw from a Bernoulli distribution with probability *p*_*i*_(*t*) determined whether delivery occurred during that interval, if it did not occur the pregnancy progressed to the next gestational week.

Once delivery occurred, pregnancies were classified into absorbing outcome states according to gestational age at delivery. Deliveries before 37 completed weeks were classified as preterm births, whereas deliveries at or beyond 37 weeks were classified as term births.

To obtain stable estimates, the simulation was repeated across a thousand Monte Carlo (random sampling) iterations, and summary measures including mean gestational duration, and cumulative incidence of preterm birth.

The temperature-attributable fraction of preterm birth was estimated by comparing the expected number of preterm births under the observed or projected temperature distribution with the expected number under a counterfactual temperature scenario (Gasparrini et al. 2014)st. The counterfactual was defined by fixing temperature exposure at the minimum predicted risk temperature (MPRT) estimated from the retrospective population, while keeping all other individual characteristics, seasonality, baseline hazard, and model coefficients unchanged. The same MPRT was applied to both the historical and projected periods to ensure comparability of attributable fractions over time.

For each pregnancy, weekly hazards between gestational weeks 22 and 36 were predicted from the fitted Cox-DLNM model under two exposure scenarios: the observed or projected temperature history and the MPRT counterfactual. Weekly hazards were accumulated across gestation and converted into the probability of preterm birth as:

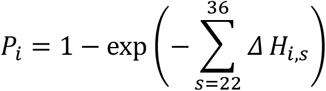

where Δ*H*_*i,s*_ represents the predicted hazard increment for pregnancy i at gestational week s.

Individual probabilities were then summed within each calendar year to obtain the expected number of preterm births under the observed or projected temperature scenario:

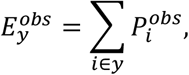

and under the MPRT counterfactual:

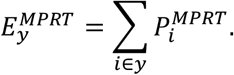

The temperature-attributable number was calculated as:

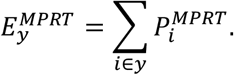

and the annual temperature-attributable fraction as:

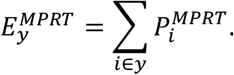

For presentation, the attributable fraction was expressed as a percentage:

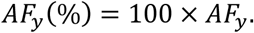

Positive values indicate a higher expected burden of preterm birth under the observed or projected temperature distribution than under the MPRT counterfactual, whereas negative values indicate a lower predicted burden relative to the counterfactual.

As an internal validation of the microsimulation framework, the simulation procedure was applied to the historical study period and simulated preterm birth estimates were compared with the corresponding observed data. Monte Carlo prediction intervals were generated from repeated simulations to assess whether the observed historical values were reproduced by the simulation framework.

## Results

### Descriptive analysis

We included 4,338,825 singleton births, of which 211,397 (4.9%) were preterm (Table 1). The median gestational age was 41 weeks (Interquartile range (IQR): 40–42) overall and among term births, compared with 36 weeks (IQR: 35–37) among preterm births.

**Table 1.** Characteristics of the study population, overall and according to preterm birth status. Values are presented as median (IQR) for continuous variables and n (%) for categorical variables. IQR is the interquartile range; n is the number; % is the subgroup percentage.

| Characteristic | Overall, N =<br>4,338,825 | Term births,<br>N = 4,127,428 | Pre-term births,<br>N = 211,397 |
| --- | --- | --- | --- |
| **Weeks in pregnancy** | 41 (40, 42) | 41 (40, 42) | 36 (35, 37) |
| **Income (for different quantiles)** |  |  |  |
| 0 | 22,326 (0.5%) | 20,661 (0.5%) | 1,665 (0.8%) |
| 1 | 505,311 (12%) | 480,602 (12%) | 24,709 (12%) |
| 2 | 826,209 (19%) | 785,558 (19%) | 40,651 (19%) |
| 3 | 939,477 (22%) | 893,784 (22%) | 45,693 (22%) |
| 4 | 995,259 (23%) | 946,882 (23%) | 48,377 (23%) |
| 5 | 1,050,243 (24%) | 999,941 (24%) | 50,302 (24%) |
| **Previous maternal hosp.** | 1,377,512 (32%) | 1,283,313 (31%) | 94,199 (45%) |
| **Age of mother** | 29.2 (25.6, 33.0) | 29.2 (25.6, 33.0) | 29.1 (25.2, 33.3) |
| **Year of birth** | 1996 (1985, 2008) | 1996 (1985, 2008) | 1995 (1985, 2007) |
| **Birth trimester** |  |  |  |
| April-June | 1,162,010 (27%) | 1,108,048 (27%) | 53,962 (26%) |
| Jan-March | 1,102,230 (25%) | 1,046,949 (25%) | 55,281 (26%) |
| July-September | 1,101,185 (25%) | 1,049,957 (25%) | 51,228 (24%) |
| October-December | 973,400 (22%) | 922,474 (22%) | 50,926 (24%) |
| **Mothers' country of birth** |  |  |  |
| Other | 722,483 (17%) | 686,488 (17%) | 35,995 (17%) |
| Swedish | 3,616,342 (83%) | 3,440,940 (83%) | 175,402 (83%) |

Maternal and pregnancy characteristics of the study population are presented in Table 1. The median maternal age was approximately 29 years, and most mothers were born in Sweden (83%). Previous maternal hospitalization was reported for 31% of term births and 45% of preterm births. Household income, which was grouped into quantiles representing increasing levels of income from the lowest to the highest category, and other maternal characteristics are also summarized in Table 1.

Births were distributed relatively evenly throughout the year, with the largest proportion occurring between April and June (27%). Among preterm births, the proportions occurring from January to March and October to December were (26%) and (24%), respectively, with the remaining births distributed across the other quarters.

### Retrospective analysis

The estimated associations between temperature and preterm birth were modest (Figure 1). Very low temperatures were associated with an increased hazard of preterm birth at short lags. The strongest association occurred at -10°C at lag 0 and weakened as the lag increased. Associations were smaller at -5°C, estimates at 0°C remained close to the null throughout the lag period.

**Figure 1.**
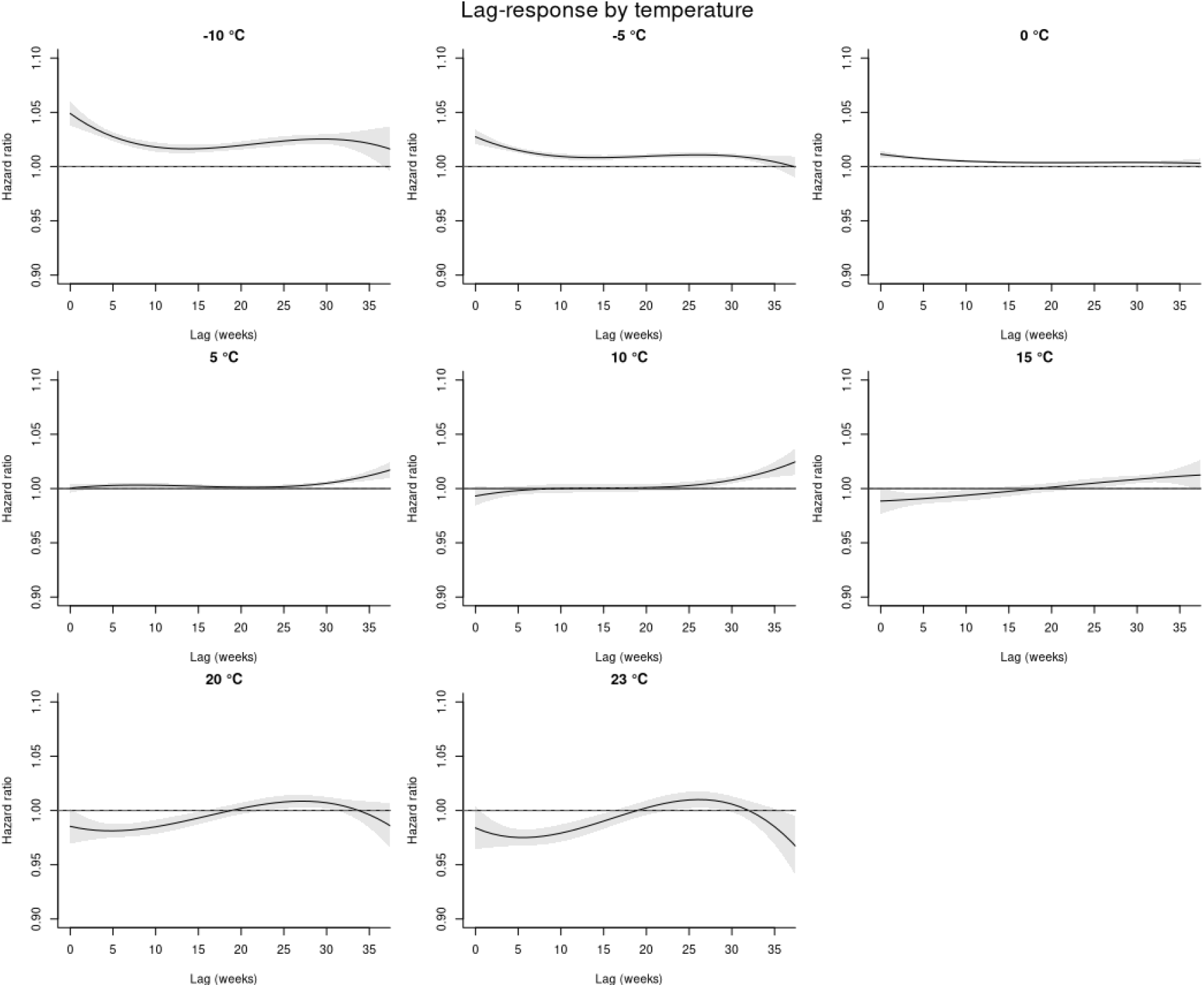
Lag-specific associations between temperature and preterm birth. The x-axis represents gestational lag in weeks, and the y-axis shows the hazard ratio 8HR) for preterm birth. Solid line indicate point estimates and shaded areas the 95% confidence intervals (95% CI)

At warmer temperatures (from 10°C to 23°C), hazard ratios were generally below or close to 1.0 at short lags and increased gradually at intermediate and longer lags. However, the magnitude of these associations remained small, and uncertainty increased towards the end of the lag period.

### Microsimulation analysis

We next examined temporal trends in three outcomes: the annual number of preterm births, the annual cumulative incidence, and the attributable fraction of preterm birth due to ambient temperature for the historical (1974-2018) and future (2025-2055) conditions using an ensemble of climate projections under SSP1-2.6 and SSP3-7.0.

The absolute number of preterm births showed a gradual increasing trend between 2025 and 2055 across the simulations (Figure 2). In contrast, the annual cumulative incidence of preterm birth remained relatively stable over the projection period, with only small differences between climate scenarios (Figure 3).

**Figure 1.**
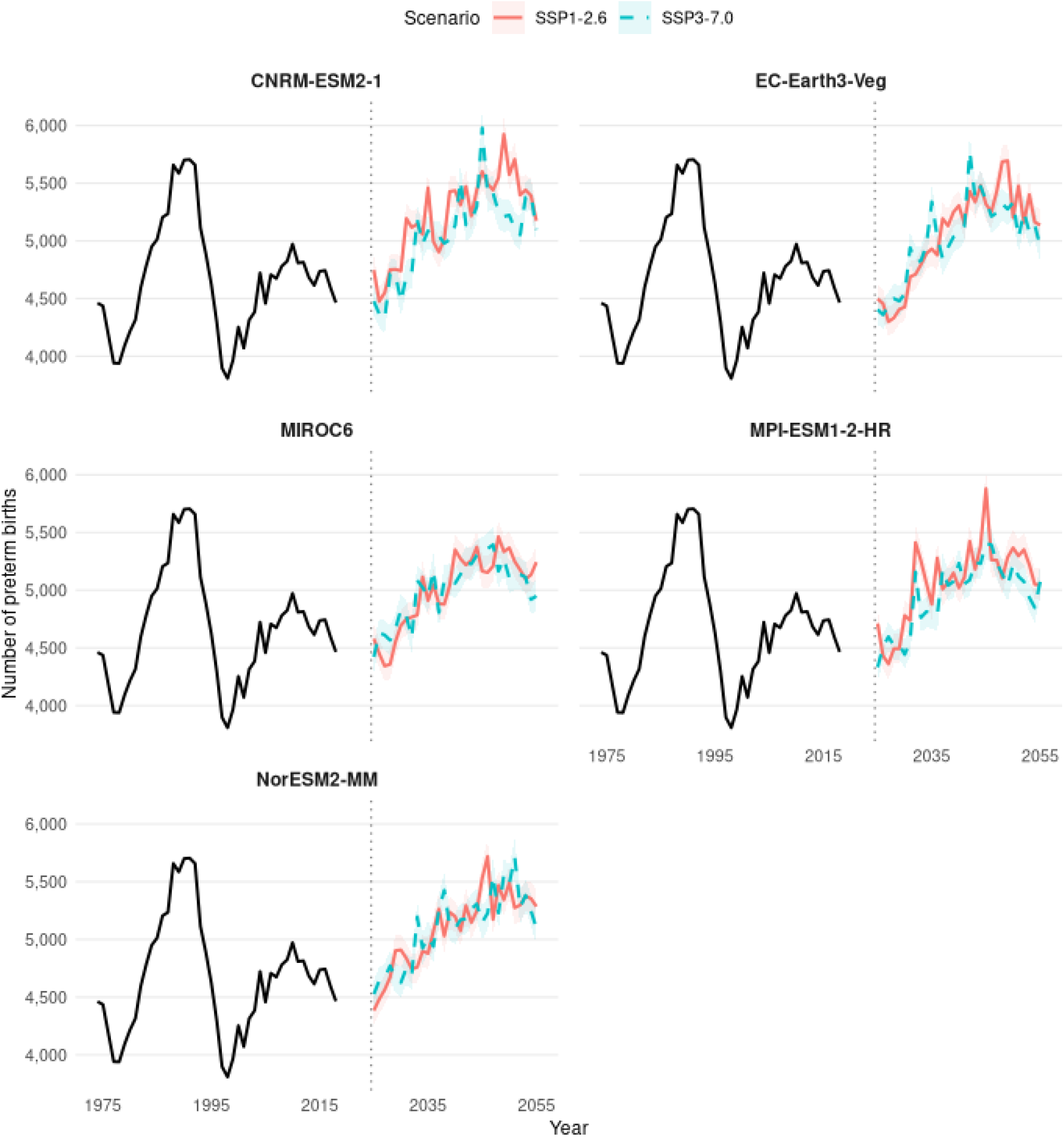
Historical and projected annual numbers of preterm births by climate model and emissions scenario. Each panel represents one global circulation model. Historical counts for 1974–2018 are shown in black and repeated across panels for reference. Projected annual counts for 2025–2055 are shown separately for the SSP1-2.6 and SSP3-7.0 scenarios; shaded bands represent the 95% Monte Carlo intervals from 1,000 stochastic realizations. The vertical dashed line separates the historical and future periods.

**Figure 3.**
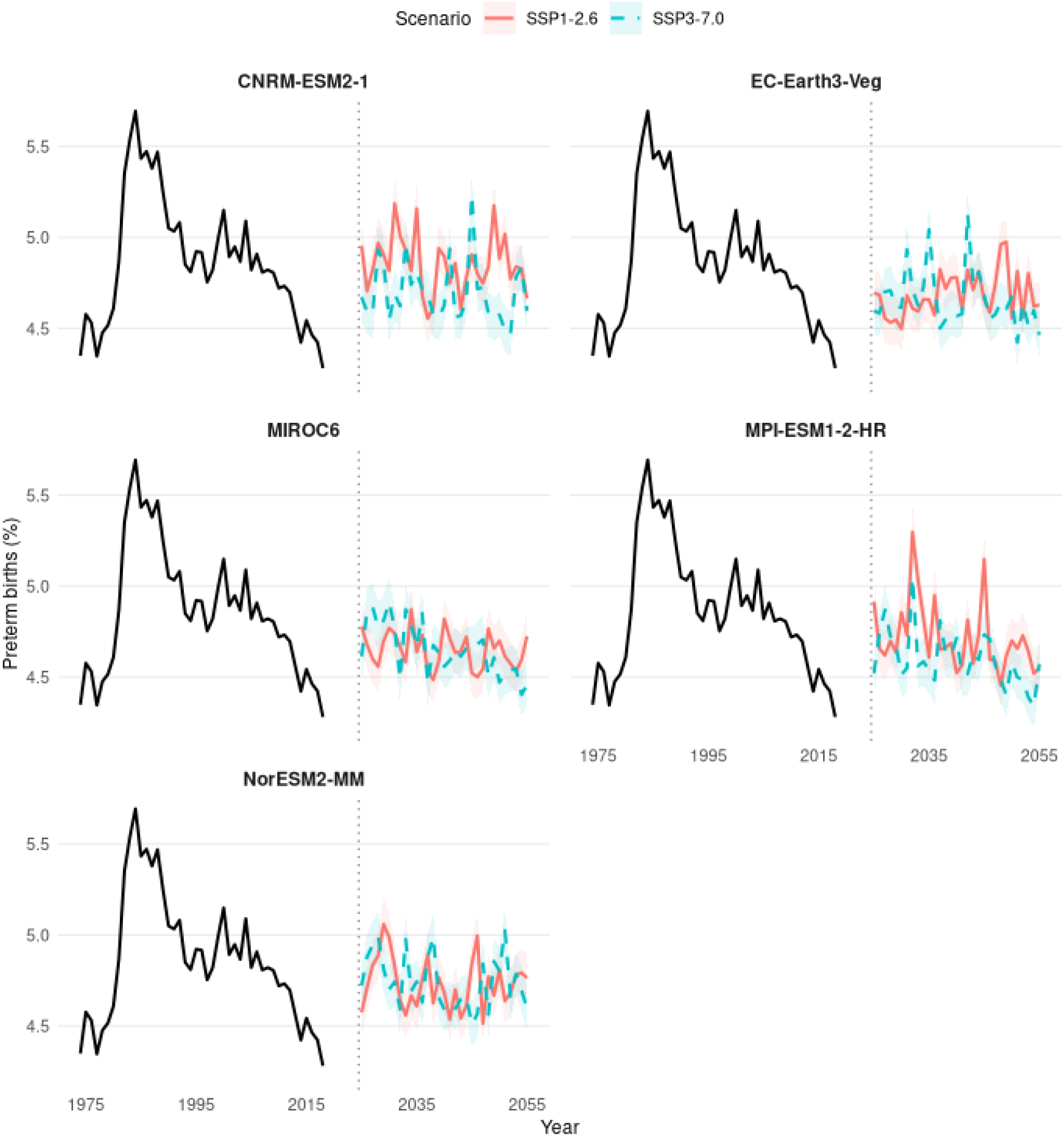
Historical and projected annual preterm birth proportion by climate model and emission scenario. Each panel represents one global circulation model. Historical observations for 1974–2018 are shown in black and are repeated across panels for reference. Projected annual proportions for 2025–2055 are shown separately for SSP1-2.6 and SSP3-7.0; shaded bands represent the 95% Monte Carlo intervals from 1,000 stochastic realizations. The vertical dashed line separates the historical and projected periods.

The annual temperature-attributable fraction of preterm birth also remained relatively stable over time across climate models and emissions scenarios, with only small year-to-year fluctuations and limited separation between projections (*Figure 4*). There was no clear temporal trend in the proportion of expected preterm births attributable to temperature between 2025 and 2055. The projected results showed broadly consistent temporal trends across different scenarios of future temperature (*Figures 2-4*). Overall, the absolute number of preterm births was projected to increase, while the annual cumulative incidence and temperature-attributable fraction remained broadly stable. This suggests that the increasing burden reflects growth in the projected number of pregnancies rather than an increase in temperature-related preterm birth risk.

**Figure 4.**
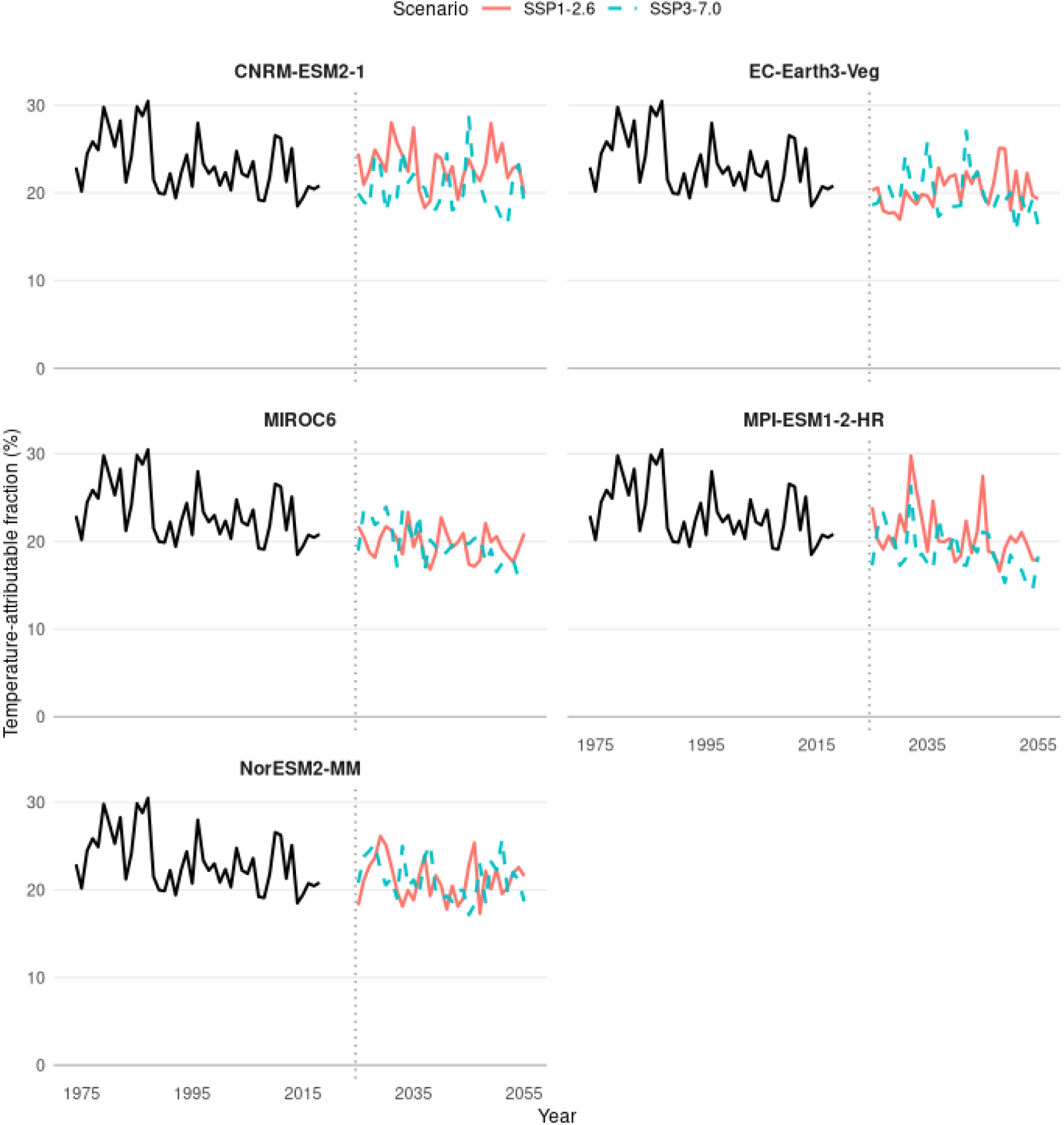
Annual temperature-attributable fraction of preterm birth from 1974 to 2055. Historical estimates are shown in black, while projected estimates for 2025–2055 are shown for each climate model under the SSP1-2.6 and SSP3-7.0 scenarios. The vertical dashed line separates the historical and future periods.

Adjustment influenced the estimated temperature–preterm birth associations, with differences in the lag–response patterns observed across temperatures (Appendix Figure 1). These differences were generally more pronounced when comparing the unadjusted model with the fully adjusted model including seasonality, suggesting that seasonal and other measured covariates contribute to the observed temperature–preterm birth relationship (Appendix Figure 2).

The estimated lag–response associations were generally robust to alternative degrees of freedom for seasonal adjustment, with only minor differences across most temperatures (Appendix Figure 3). Greater variation between specifications was observed at warmer temperatures, particularly at 20°C and 23°C, suggesting that estimates at the upper end of the temperature distribution were somewhat more sensitive to the choice of seasonal adjustment (Appendix Figure 4).

The microsimulation framework reproduced the historical data reasonably well, with the observed preterm birth estimates falling within the Monte Carlo prediction intervals for most of the study period (Appendix Figure 5).

## Discussion

In this nationwide cohort of 4,338,825 singleton births, 211,397 (4.9%) were preterm. The retrospective analysis showed that the association between temperature and preterm birth was strongest at the coldest temperatures and at short lags, whereas higher temperatures were not associated with a clear increase in risk. In the climate-storyline analysis, the historical exposure-response relationship was incorporated into individual-based microsimulations using five climate models and two emissions scenarios. From 2025 to 2055, the annual cumulative incidence of preterm birth and the temperature-attributable fraction remained broadly stable across storylines, whereas the absolute number of preterm births increased. This increase was driven mainly by growth in the projected number of pregnancies rather than by an increase in temperature-associated risk.

Our findings in Sweden are consistent with previous evidence that cold exposure may increase the risk of preterm birth. A study in Guangzhou found a U-shaped relationship with stronger associations for cold than for heat, and a nationwide Chinese study associated cold spells shortly before delivery with increased risk but found no stable association with heatwaves (He et al., 2016; Yu et al., 2023). A recent meta-analysis likewise found increased short-term risks following both extreme cold and heat (Khosravipour and Golbabaei, 2024). The absence of a clear heat association in our study may reflect Sweden’s comparatively moderate upper temperature range and the use of weekly mean temperature, which may mask the effect of short heat episodes.

The limited change in projected preterm birth incidence is consistent with the modest temperature-preterm birth associations observed in the retrospective analysis. Although temperatures is expected to increase over future decades in Sweden, much of this warming occurred within a range of the temperature distribution where the estimated exposure-response relationship was relatively flat. Meanwhile, cold exposure, the main temperature-related risk identified in our analysis, is projected to remain relatively stable in Sweden. As a result, changes in temperature translated into only small changes in individual preterm birth risk, leading to little variation in the projected incidence over time. A similar pattern was observed for the temperature-attributable fraction, suggesting that the proportion of preterm births associated with temperatures away from the minimum predicted risk temperature (MPRT) changed only modestly across the projection period. However, a relatively unchanged attributable fraction does not imply a stable absolute burden. If the number of pregnancies increases, the number of attributable preterm births may also rise even if the attributable fraction changes little. Distinguishing between relative and absolute burden is therefore important when interpreting future health impacts, as modest changes in risk can still correspond to a larger number of affected pregnancies in a growing population.

This study has several important strengths. The analysis was based on a nationwide population-based cohort with individual-level information, providing substantial statistical power and reducing the likelihood of selection bias. The microsimulation estimated risk separately for each pregnancy using maternal characteristics, gestational timing, parish of residence, and weekly temperature history rather than applying a single average effect. This preserved individual heterogeneity while translating the historical exposure–response relationship into projected population burden. An additional strength was the use of multiple climate models and emissions scenarios, allowing the robustness of the projected patterns to be assessed across alternative plausible future temperature trajectories rather than relying on a single climate projection.

This study has also potential limitations. Parish-level weekly mean temperature is a proxy for personal exposure and may not capture indoor conditions or brief temperature extremes, potentially attenuating the estimated associations. The projections also assume that the historical temperature–preterm birth relationship remains applicable through 2055, although changes in adaptation, housing, healthcare, behaviour, and population vulnerability could modify this relationship. Furthermore, the Monte Carlo intervals represent stochastic variation within the simulations but do not capture the full uncertainty in the exposure–response estimates, climate projections, or future pregnancy population. Finally, the analysis estimates only the temperature-related component of climate risk and does not include other pathways, such as air pollution, wildfire smoke, flooding, or infectious disease.

These findings have implications for climate adaptation and maternity-service planning. The cold-related association suggests that protection of pregnant women from low temperatures remains relevant in Sweden, even in a warming cliamte. At the same time, continued surveillance of heat-related pregnancy outcomes is warranted because future heat extremes could exceed the range represented in the historical data. For healthcare planning, stable preterm-birth incidence should not be interpreted as stable service demand: a growing number of pregnancies could increase the absolute need for obstetric and neonatal care even without an increase in temperature-associated risk. Climate-health assessments should therefore report both relative and absolute burden and incorporate demographic projections when planning future services.

## Conclusion

In conclusion, the retrospective analysis identified the clearest increase in preterm-birth risk at the coldest temperatures and short lags, with no clear increase at higher temperatures. When the historical exposure-response relationship was applied across multiple future climate storylines, preterm-birth incidence and the temperature-attributable fraction changed little through 2055, whereas the absolute number of preterm births increased. Under the assumptions of the model, this increase was driven mainly by growth in the number of pregnancies rather than by increasing temperature-associated risk.

## Data Availability

The registry data used in this study are not publicly available. Access to the data is subject to approval by the relevant data holders and requires appropriate ethical and data-access permissions.

## Funding

This Perspective was supported by the Swedish Research Council for Sustainable Development (Formas; 2022-01845 and 2023-01774), the Swedish Research Council for Health, Working Life and Welfare (2024-00833), and the Swedish Research Council (2025-02804 and 2022-06599), Karolinska Institutet (2022-00017 and 2025-01132).

## Ethical approval

The Swedish Ethical Review Authority approved studies (reference 2020-05516, 2022-05648-02 and 2022-02626-01) to access de-identified, whole-population registry data, collected without individual consent from routine healthcare records and administrative registers.

## Appendix

**Appendix Figure 1:**
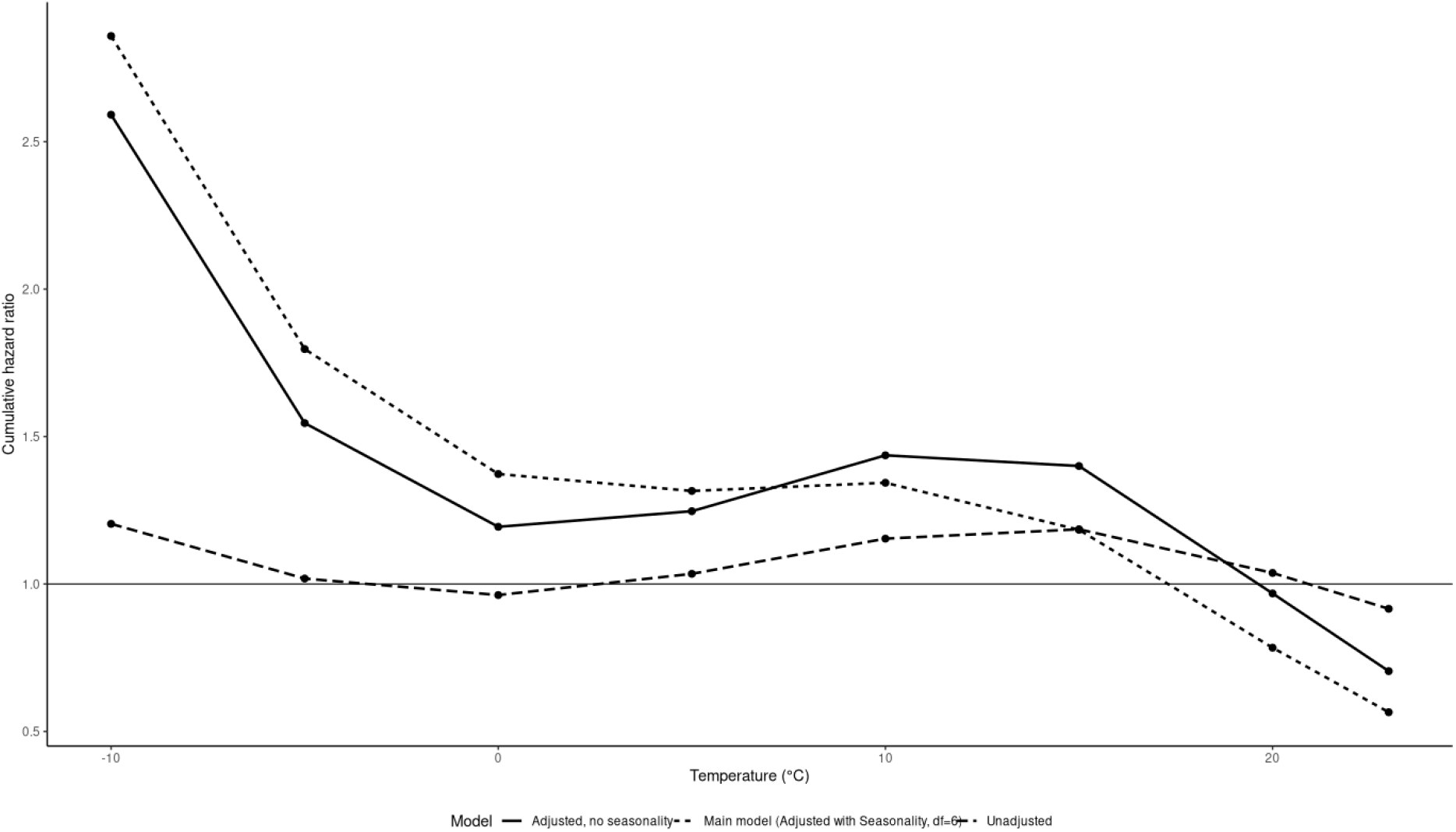
Overall cumulative temperature–preterm birth hazards comparing the primary model with the adjusted model without seasonal adjustment and the unadjusted temperature-only model. Hazard ratios are relative to 10°C.

**Appendix Figure 2:**
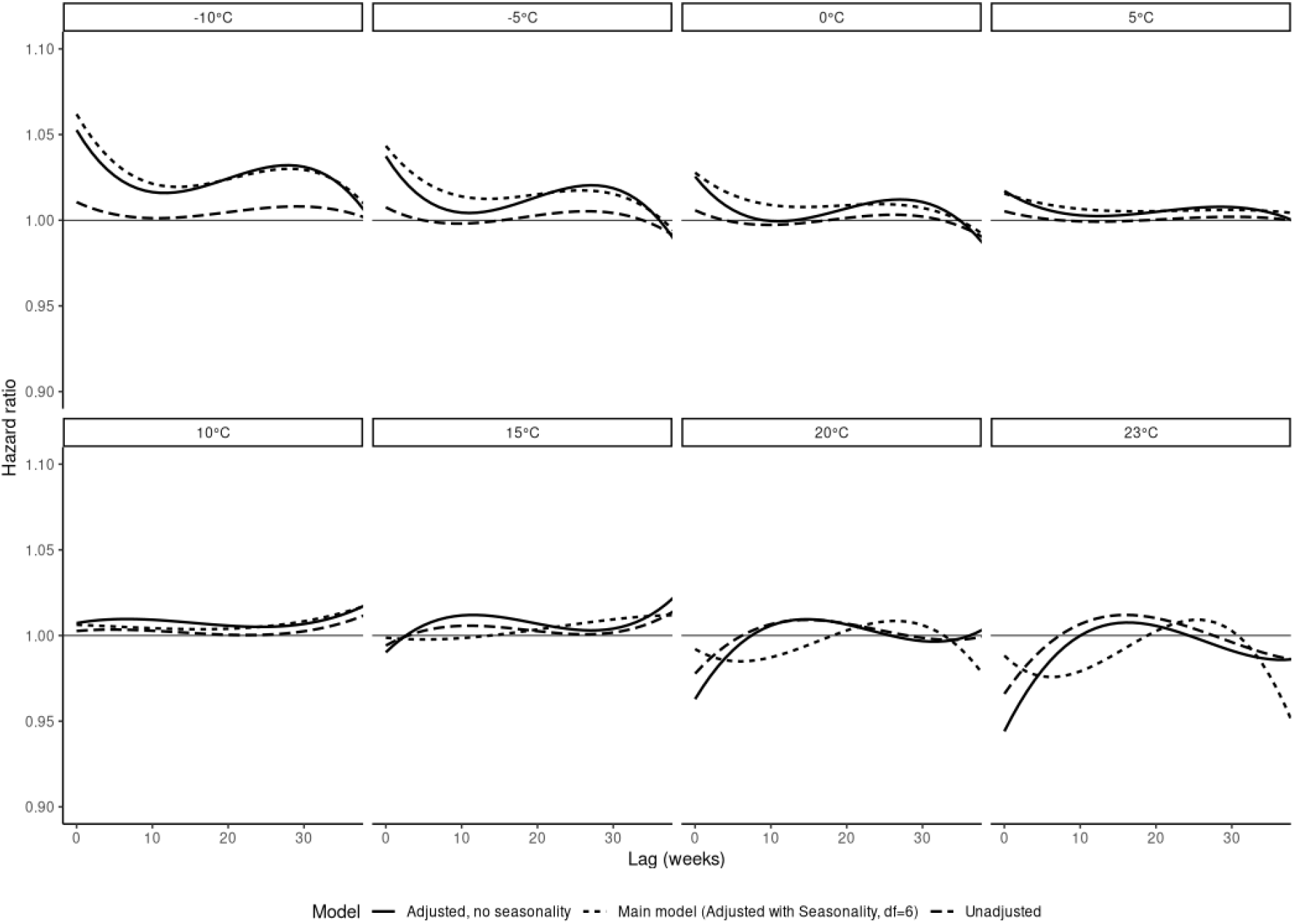
Lag-specific hazard ratios for selected temperatures comparing the primary model with the adjusted model without seasonal adjustment and the unadjusted temperature-only model. Hazard ratios are relative to 10°C.

**Appendix Figure 32:**
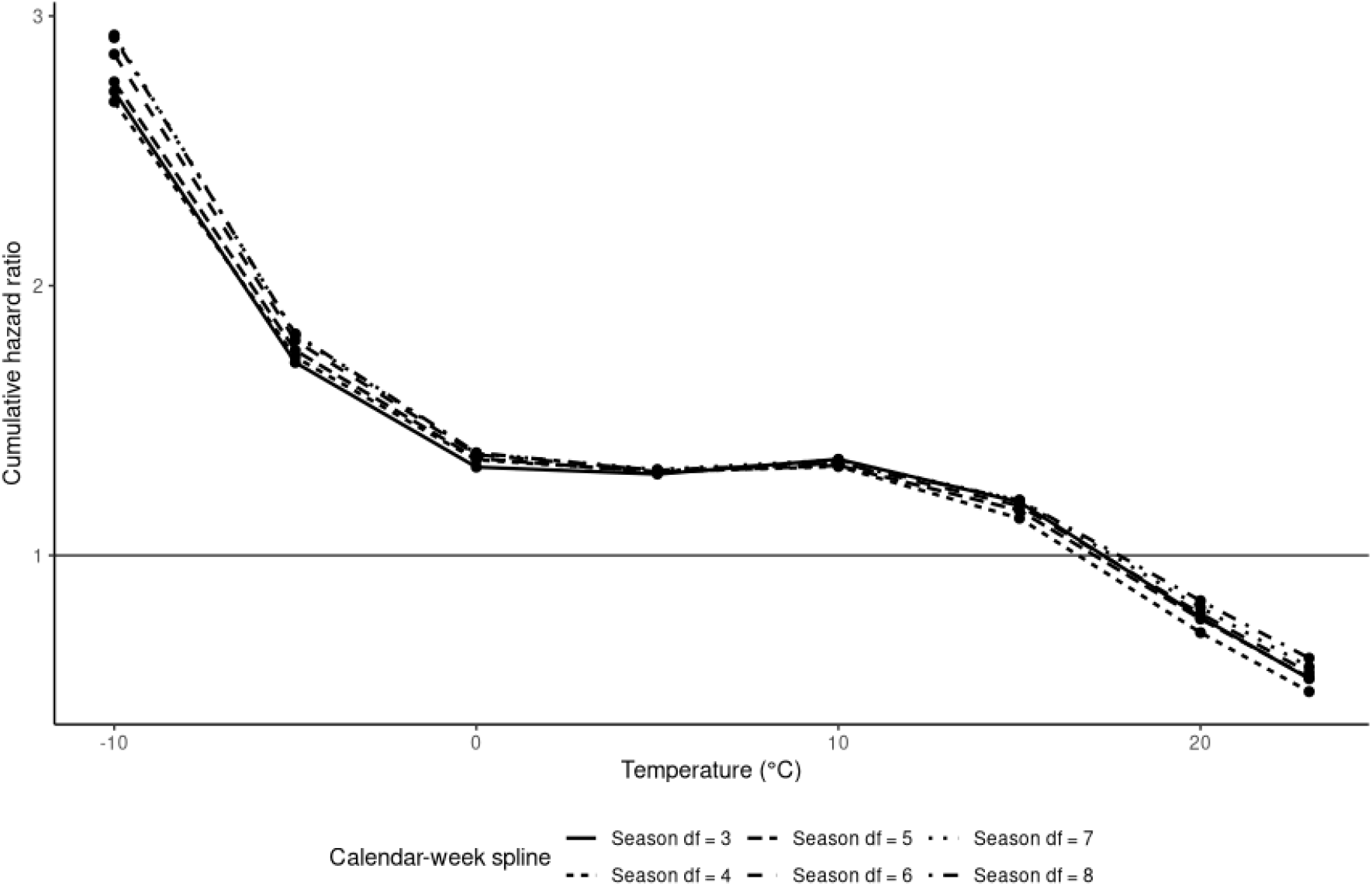
Overall cumulative temperature–preterm birth associations using alternative degrees of freedom for calendar-week seasonality. The primary model used 6 degrees of freedom. Hazard ratios are relative to 10°C.

**Appendix Figure 4:**
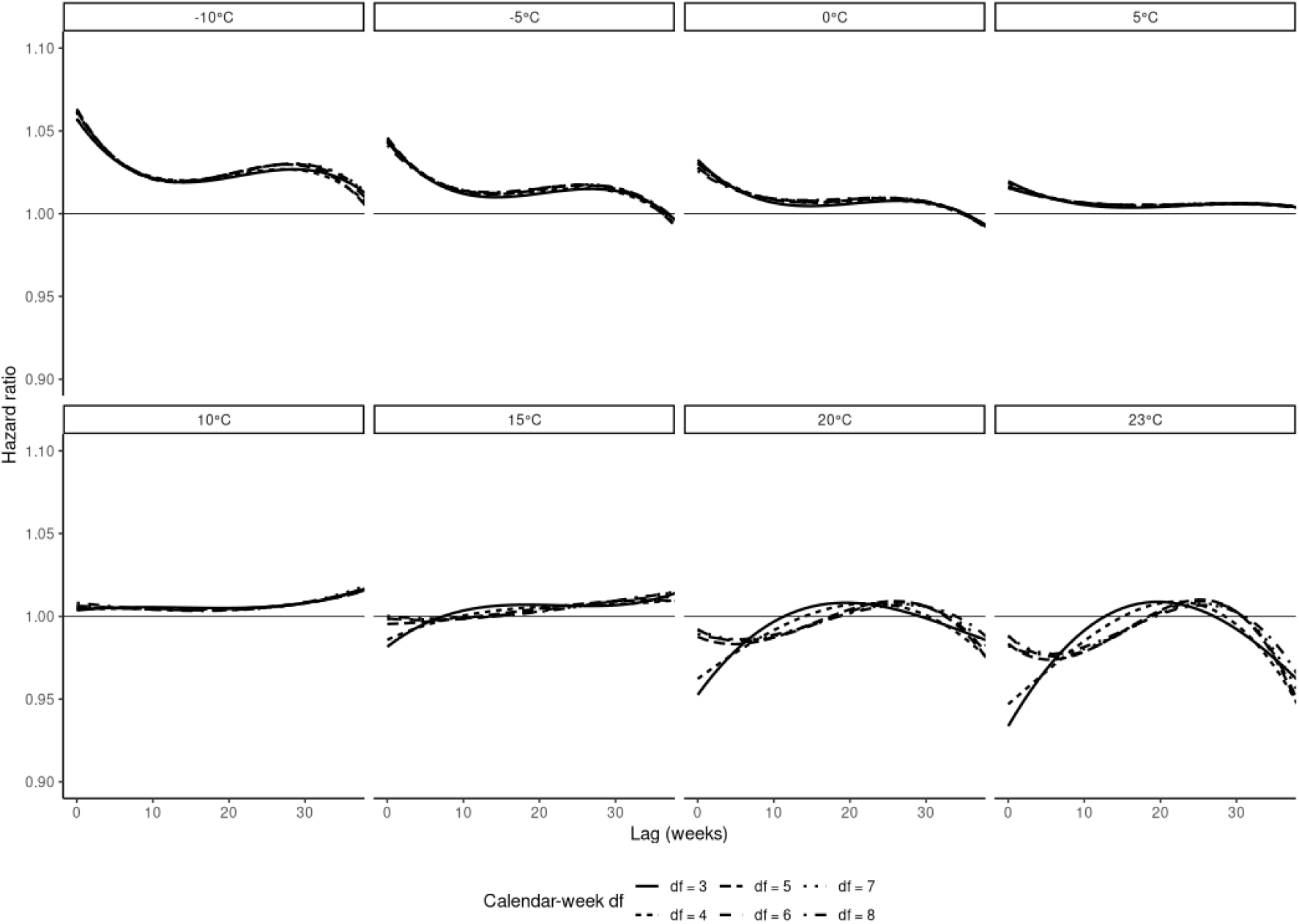
Lag-specific hazard ratios for selected temperatures using alternative degrees of freedom for calendar-week seasonality. The primary model used 6 degrees of freedom. Hazard ratios are relative to 10°C.

**Appendix Figure 5:**
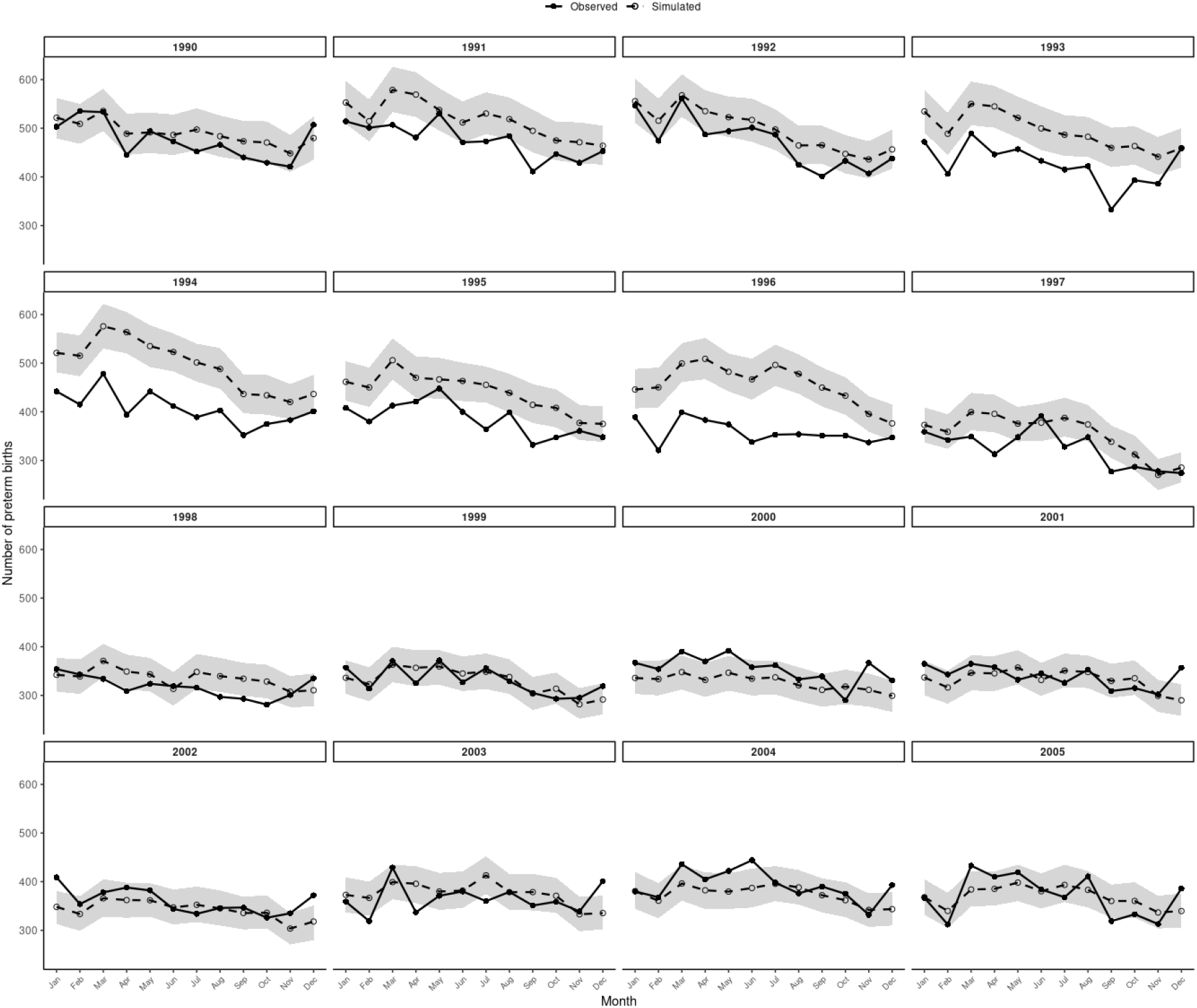
Observed and simulated monthly numbers of preterm births by calendar year, 1990–2005. The shaded area represents the 2.5th–97.5th percentile across 1,000 Monte Carlo simulations.

## Notes

### Competing Interest Statement

The authors have declared no competing interest.

